# Optical imaging of micro-droplets of dried saliva for oral squamous cell carcinoma diagnosis

**DOI:** 10.1101/2024.09.27.24314396

**Authors:** Letícia Foiani, Gabrielle Nepomuceno, Julia Toledo, Mariana Alves, Nayara Rodrigues, Celso Bandeira, Mônica Alves, Janete Almeida, Herculano Martinho

## Abstract

Oral cancer, the sixth most common worldwide, is often diagnosed at an advanced stage, impacting patient survival and mortality. Liquid biopsy offers the potential for cancer diagnosis, enabling dynamic tumor monitoring and disease surveillance. Here we validates a novel diagnostic approach using optical images of dried micro-droplets (volume of one ***µ***l) of saliva samples on glass and platinum substrates, employing Logistic Regression and Support Vector Machine (SVM) models. For each model, accuracy, sensitivity, specificity, and area under the ROC curve were calculated. Our findings indicated that optical density and surface area (SA) obtained from optical images of microdroplets are suitable paramters of discriminating oral cavity squamous cell carcinoma and health individuals. SVM models demonstrate impressive accuracy of 88.10% on glass and 95.00% on Pt substrates, ensuring robust and accurate detection of oral cancer based on these salient features.

## 1 Introduction

Oral cancer is a malignant tumor that arises from the oral cavity and lips as one of the most common type of head and neck cancer [1, 2]. In 90% of cases, oral cancer originates from squamous cells. Therefore it is called squamous cell carcinoma, or oral cavity squamous cell carcinoma (OSCC) [2]. The annual incidence of OSCC worldwide in 2020 was 377,713 cases for both sexes, with 177,757 deaths. It represents the 8^*th*^ type of cancer in men according to the data from the Global Cancer Observatory[3].

Several investigations pointed out that the use of tobacco (including smokeless tobacco). the excessive consumption of alcohol, the unprotected sun exposure, and even the lack of mouth hygiene, as the main routes leading to oral cancer.[4–6] According to Reidy and Stassen[7] smoking can contribute to the development of OSCC due to the changes caused in the cellular structure of the oral mucosa, contributing to the deficiency in immune system which provides conditions for the development of an environment favorable to malignant transformation.

Early diagnosis is an important target, since a significant cause of the increasing number of OSCC cases is related to limitations in early and accurate diagnosis of the disease [2, 8, 9].

In general, the majority of OSCC diagnoses are made when the disease is at an advanced level, which decreases survival and increases mortality for patients. Histopathological morphological analysis performed on a biopsed tissue is the gold standart for cancer, OSCC in particular, diagnosis. However, this method has some drawbacks such as the limitation of the region to be analyzed within a tumor which has high heterogeneity. The method is performed by visual inspection by a trained professional, which frequently leads to interpretative conflicts. Conducting large-scale population screening and patient monitoring is hindered by various challenges, including invasiveness, high costs, time consumption, and the requirement for specialized personnel and equipment [10–12], besides to contraindications associated with systemic diseases [13].

Liquid biopsy is a potentially useful technique for the characterization of samples for cancer diagnosis based on body fluids. It enables monitoring of dynamic tumor burden and active surveillance of disease recurrence [14]. The identification of new molecular biomarkers and the development of new technologies capable real time sensing at the lowest possible costs are an urgent need enabling early diagnosis, as well as the development of drugs with appropriately chosen for each individual [11, 15]

Salivary fluid has several practical advantages among other body fluids for biomedical determinations. Its collection is non-invasive and painless, without discomfort to patients. In addition, salivary fluid collection tends to be a more economical procedure, since it is easily obtained, transported, and stored[16]. Investigations have shown that in saliva biomarkers for oral diseases as, e.g., those associated with systemic problems can be found. Increasingly, disease-specific molecules such as breast cancer, cardiovascular disease, and human immunodeficiency virus (HIV) are being identified in this type of body fluid [17, 18].

There is great interest in the search for new technologies that allow rapid, non-invasive, and low-cost characterization of biological fluids. This need is most evident in the biomedical field, where rapid diagnosis combined with low cost is a decisive factor in reducing mortality [19]. Photonics technologies have been employed for diagnostic purposes quite promisingly to, e.g., provide early diagnosis of cancers[20], real-time evaluation of sepsis[21] and some other purposes[19, 22, 23].

Here we present an approach based on analysis of optical images of micro-droplets of saliva of healthy and OSCC individuals which enable discriminate these groups of patients.

## 2 Methodology

### 2.1 Sample collection and storage

The research was approved by the Research Ethics Committee of ICT-UNESP (CAAE 42387315.0.0000.0077). After being informed about the proposition and conditions of this study, patients who agreed to participate in the research signed the Informed Consent Form. Non-inclusion criteria were patients who had already undergone any type of oncological treatment, whether surgery, radio, or chemotherapy in any organ or system, as well as cases of lip cell carcinoma.

Samples consisted of saliva from consecutive patients who were admitted for diagnosis and treatment of OSCC at the Head and Neck Surgery Service of the Municipal Hospital José Carvalho de Florence in the city of São José dos Campos/SP - Brazil, the Maternity and Hospital Celso Pierro of the Pontific Catholic University of Campinas/SP - Brazil (PUC-Campinas) and the Municipal Hospital Mário Gatti of Campinas/SP, with a diagnosis of OSCC aged over 18 years. After collection, samples were immediately stored in Nalgene R cryogenic tubes, properly identified and numbered, and kept under refrigeration in liquid nitrogen during transportation. Afterwards, they were immediately stored in an ultrafreezer (−80^*°*^C) of Thermo Fisher Scientific Inc. Samples were separated into two groups labeled CA (patients with OSCC) and CN (control group).

For each group, 21 samples were used in triplicate (generating 63 droplets for each group) for every test conducted here.

### 2.2 Sample preparation conditions and optical imaging

The main parameters to be considered to ensure reproducibility in drying procedure were sample dilution, drying temperature, and relative humidity during drying[24]. The dilutions tested were 1:2; 1:5; 1:10; 1:15; 1:20; 1:30; 1:35, in addition to the undiluted sample. These dilutions were tested based on results reported in the literature[24].

Published work involving other biofluids[24] indicates that the best condition involves drying in an environment with 70% relative humidity. Thus, to ensure the conditions for saliva preparation, a stable environment was created with Sodium Chloride solution that was placed inside the desiccator and maintained in equilibrium for 24 hours, allowing for a relative humidity of 80%. The temperature of the environment was controlled at 20°C.

After stabilization of the system, samples were removed from the ultrafreezer, thawed at room temperature at 25°C, and then 1*µ*L of each pipetted onto the substrates. The drops were prepared in triplicate. Substrates containing saliva were then placed in the desiccator for relative-humidity controlled drying as described above. We tested 3.5 inches circular Pt substrates and 20*×*20*×*0.16 mm glass coverslips. Figure 1 shows how the samples were arranged in each substrate. Optical images were taken using a Zeiss Axio Observer microscope.

**Fig. 1.**
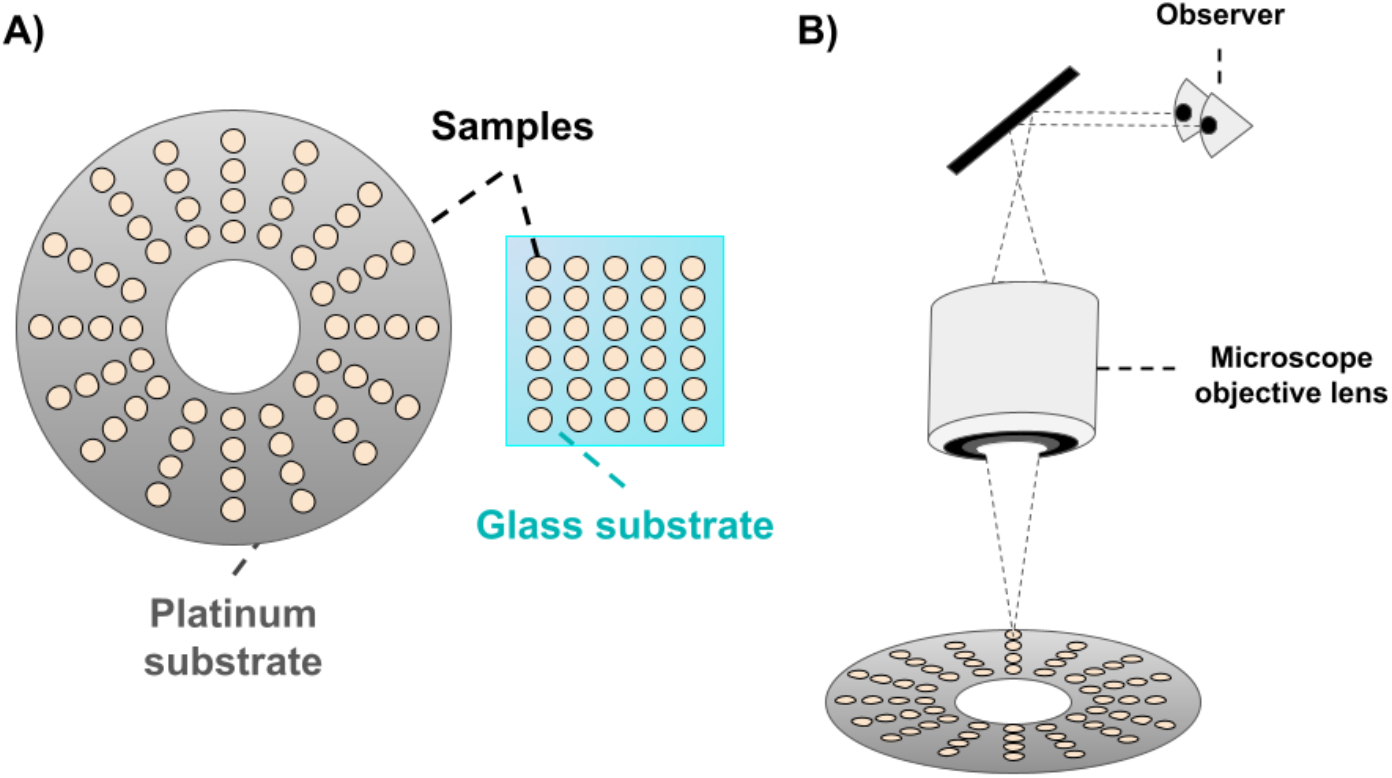
Arrangement of microdroplets of saliva samples on Pt and glass substrates (A). Each dried microdroplet was imaged through an optical microscope (B).

### 2.3 Atomic force microscopy

Atomic Force Microscopy (AFM) imaging was performed on the Pt substrate used for pipetting samples. The results of the AFM analysis unveiled characteristic grooves on the substrate surface. These grooves presented an average depth of approximately 245 *±* 88 nm. On the other hand glass covership presented a roghness of 5 *±* 2 nm.

### 2.4 Data analysis

Optical images were treated and analyzed using the ImageJ package[25]. Here we conducted a meticulous statistical analysis of the samples imaging employing two machine learning techniques, specifically logistic regression (LR) and support vector machine (SVM). LR was used to scrutinize a dataset comprising one or more independent predictior variables that determine an outcome. Usually dealing with binary classification problems, LR is based on modeling the logarithm of odds ratio of outcome probability *p* as function of linear combination of independent predictor (*pred*_*i*_),[26]

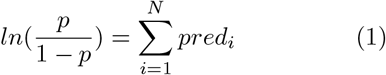

SVM is known for their robust supervised learning capabilities in classification and regression analysis, effectively segregate data points by constructing a decision boundary that maximizes the margin between different classes. This feature renders them exceptionally valuable for dissecting intricate datasets with distinct margins of separation. Integrating both LR and SVM in the study aimed to leverage the individual strengths of each technique, resulting in a more comprehensive and robust analysis of the samples, consequently enhancing the accuracy and dependability of the findings. [27–29] In detail, SVM constructs a hyperplane or set of hyperplanes in a high dimensional space constituted by preditor variables, which is employed for classification purposes. Separation is achieved by the hyperplane that has the largest distance to the nearest training-data point of any class.[30]

Performance metrics were computed for each algorithm, including accuracy, specificity, sensitivity, and the Receiver Operating Characteristic (ROC) curve. Accuracy serves as an indicator of the overall correctness of the classification model, while specificity and sensitivity gauge the model’s ability to accurately identify negative and positive instances, respectively. The ROC curve, a graphical representation illustrating the performance of a classification model at various thresholds, further enriched the analysis. The integration of these statistical methods and performance metrics ensured a comprehensive and meticulous examination of the dataset, thereby bolstering the robustness and accuracy of the study’s conclusions. [31–33]

All the statistic tests and models were calculated using RStudio[34]. Graphs and figures were made using QtiPlot[35] and GIMP [36].

## 3 Results

The first part of the conducted investigation relied on dilutions at various ratios. analyzed on the hydrophobic Pt substrate in order to just find the dilution proportion that would present the greatest homogeneity of solid particles in the samples regarding their area. Representative obtained images for the various solutions are shown in Fig. 2A). Our findings indicated that increasing saliva sample concentration generates a smaller dispersion of spots and granulates on droplet overall area. This fact can be evidenced inspecting gray-scale histograms of each image (Fig. 2B). The histogram widening is evident when comparing the most diluted sample (1:35) to the most concentrated one (no dilution). Dilutions 1:35 to 1:15 presented gray-scale distribution with 3 maximums, indicating 3 possible distributing of concentration gradient inside the droplet. For 1:10 case, these 3 distributions start to overlap in 2, which in discernible in 1:5 and 1:2 cases. For pure salive a clear distribution centered on 130 grayscale value appear beyond the 80. In this sense, the 1:5 presented higher homogeneity.

**Fig. 2.**
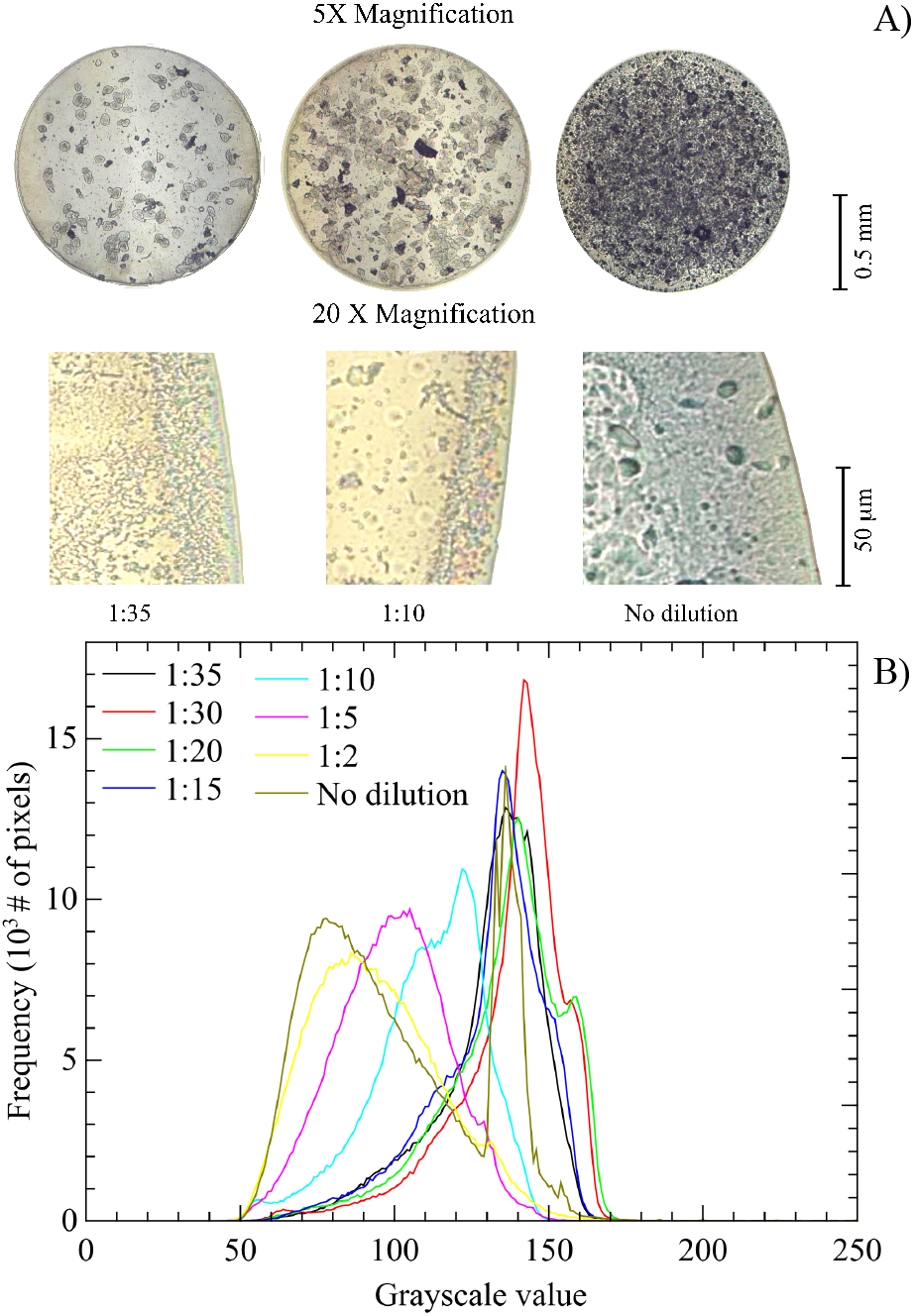
A) Some selected optical images of microdroplets of saliva for control group in 1:35 and 1:10 dilutions and no dilution as well for 5X and 20 X magnification. B) Grayscale histograms of optical images (5X magnification) of microdroplets of saliva for control group with varying dilution factors.

Another aspect to be investigated is the concentration gradient at edges of each micro-droplet (called the coffee ring effect), which occurs due to an accumulation of solid particles at the edge of the dryed drop which leaves to increasing heterogeneity. Figure 2A) shows representative images at 20X magnification. We notice the absence of coffee ring for microdroplets with concentration higher than 1:5.

Based on all the above data, it was determined that samples would be analyzed without any dilution, since it presented appreciable gray-scale factors along all the droplet size and absence of coffee ring effect. Figure 3 presents selected optical images at 5x magnification for saliva from CN and CA groups,without dilution for Pt and glass substrates. The visual inspection indicated key differences between CN and CA saliva samples. The former group usually shows greater homogeneity in the overall area of the droplet while the last one has characteristic cracks, valleys, and roughness. These qualitative characteristics can be objectively analyzed by comparing the average gray-scale histograms of all images for the CA and CN groups. Figures 3 B) and C) show the mean histograms and the respective standard deviations for both groups. The general behavior of the curves is quite distinct for the different substrates used, presenting some points that can help in discriminating CA and CN groups, especially at the extremes of the histograms (grey-scale values below 20 and above 230, for example).

**Fig. 3.**
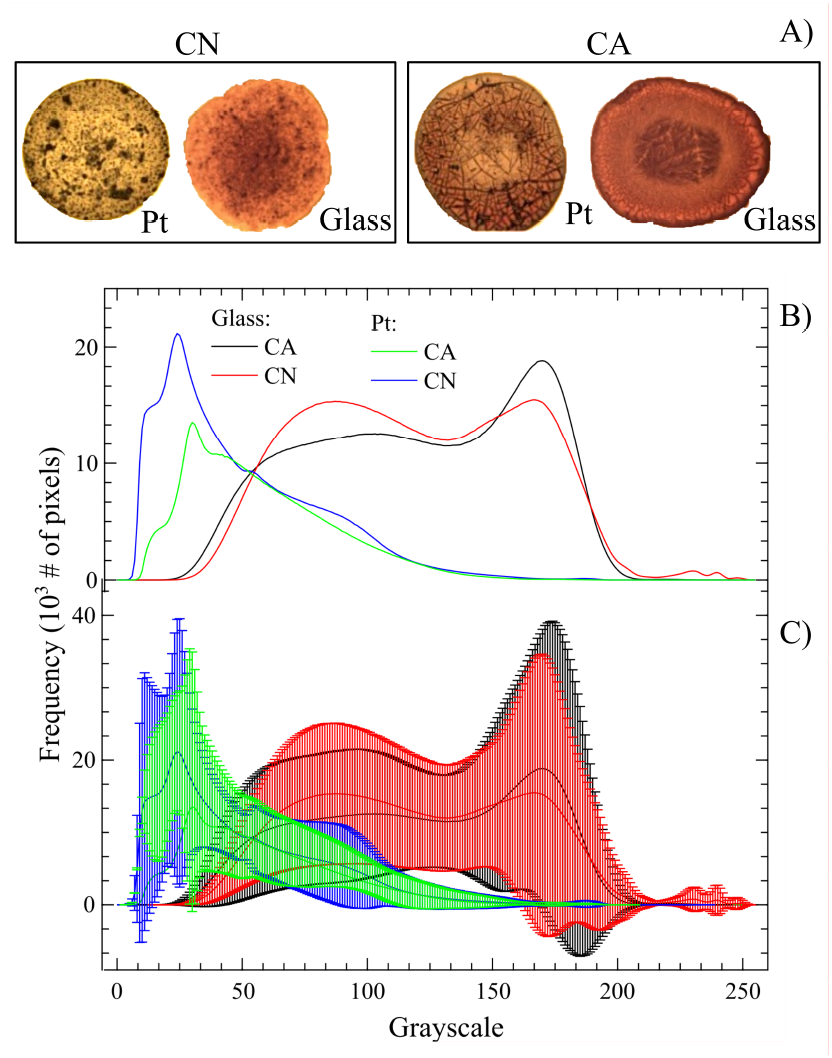
A) Selected optical images of undiluted saliva microdroplets from control (CN) and oral cavity squamous cell carcinoma (CA) groups pipetted onto Pt and glass substrates for 5X magnification. B)Average grayscale histograms of dried microdroplets of saliva of control (CN) and oral cavity squamous cell carcinoma (CA) samples analyzed on glass and Pt substrate. The corresponding standard deviation of the data are also shown (C).

The optical density (OD) parameter[37] of each microdroplet was measured, compared, and used to create mathematical models that described the samples and could help in the discrimination between the two groups. The OD values reflect the light absorption or transmission properties of the samples, and these differences might be linked to variations in sample composition, thickness, or interactions with the substrates, ultimately influencing light scattering or absorption. In addition, the surface area (SA) of each microdroplet was also analyzed. We notice that distinctive characteristics were observed in OD and SA for experimental groups and for each substrate. The differences in SA between the glass and Pt substrates could be attributed to varying sample deposition or spreading behaviors, likely arising from the distinct physical properties of the substrates. Factors such as surface roughness, hydrophicity, and surface charge of the glass and Pt may interact differently with the saliva samples, leading to discernible differences in the measured area. Additionally, the observed variations in OD among the sample groups further contributes to the substrate-dependent differences.

Statistical machine learning models based on LR and SVM were developed for the analyzes on OD and SA on different substrates. In Fig. 4, both LR and SVM models are graphically displayed. To validate the classification accuracy for each model a comparative analysis based on the area under the ROC curve (AUC) was performed. This is an evaluational metric used to verify the real performance of a given classificatory model, such as LR. Figure 5 shows the plotted graphics. All the values obtained from the above-mentioned statistical analyses are arranged in Table 1.

**Table 1.** Statistical analysis results.

| Model | Substrate | Sensitivity | Specificity | Accuracy | AUC |
| --- | --- | --- | --- | --- | --- |
| LR | Glass | 87.3 | 76.2 | 81.8 | 0.897 |
| LR | Pt | 69.5 | 62.3 | 65.8 | 0.706 |
| SVM | Glass | 80.9 | 95.2 | 88.1 | 0.880 |
| SVM | Pt | 98.3 | 91.6 | 95.0 | 0.950 |

**Fig. 4.**
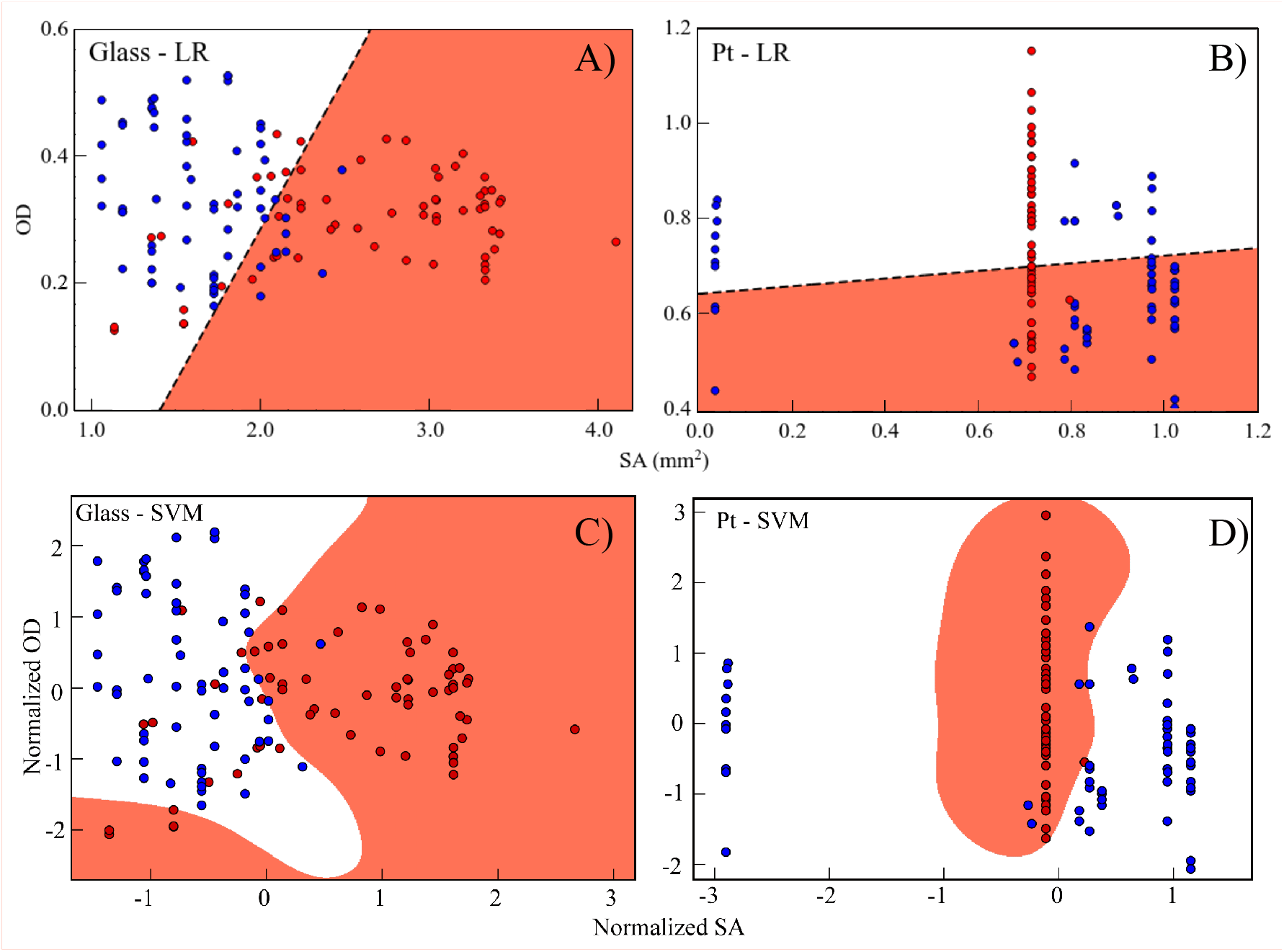
Optical density (OD) versus surface area (SA) for control (blue) and OSCC (red) groups. Straight lines at A) and represents the *p* = 0.50 logistic regression (LR) for glass (*−* 5.286*OD* + 4.25410^*−*7^*SA* + 3.390 = 0) and Pt (6.923*OD−* 3.33310^*−*6^*SA* + 4.682 = 0), respectively. Suport vector machine (SVM) models obtained for glass and Pt substrates are shown on C) and D). The red filled area in all graphs represents the disease region domain.

**Fig. 5.**
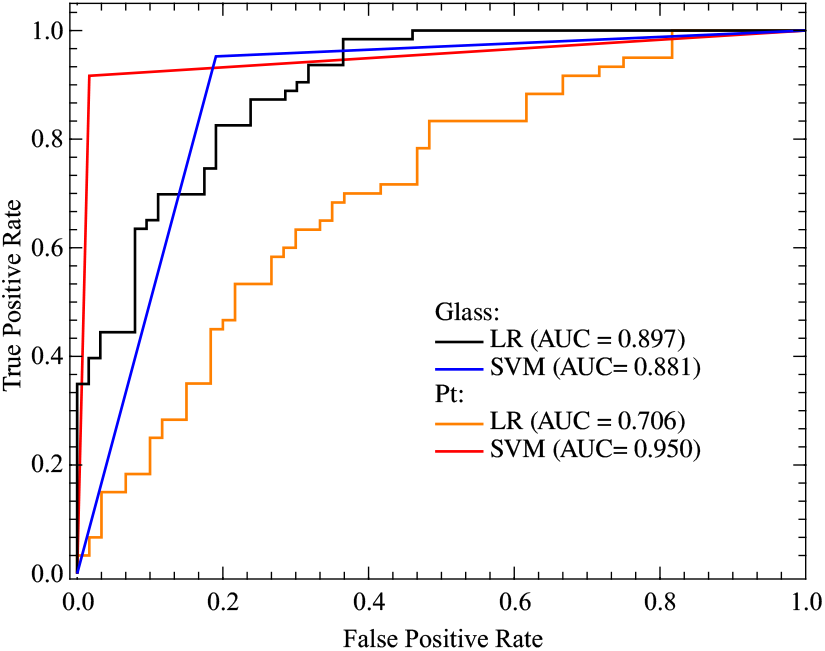
ROC curves for glass and Pt substrates in logistic regressio (LR) and suport vector machine (SVM) models. The area under ROC curve (AUC) is presented for each case.

The results obtained with LR for glass substrate indicate its high discriminative power.

Besides having an accuracy of almost 82%, it presented an AUC value higher than excelent level (AUC *>* 0.80) to validate new diagnostic ways, while sensitivity and specificity values were 87.2% and 76.2%, respectively. In addition, the SVM models applied to the glass substrate also demonstrate promising results. The SVM model achieved an accuracy of 88.1% and an AUC value of 0.880, indicating a strong predictive power and generalization capability. The sensitivity value of 80.9% and specificity value of 95.2% further underscore the model’s ability to correctly classify positive and negative cases.

The results achieved with the LR model calculated for the analyses performed on the Pt substrate show a significant difference from those previously mentioned. Besides presenting a lower accuracy than the model generated for glass, 65.83%, it presents AUC values, 0.63, and sensitivity and specificity, respectively 0.695 and 0.623, also comparatively lower, representating an poor discriminative capability in thes cases. In this subdtrate, the SVM model presented superior performance compared to the LR one, achieving an excellent accuracy of 95.00% and an AUC value of 0.950, which indicates a robust and accurate classification ability in an excellent level. The sensitivity value of 98.3% and specificity value of 91.6% highlight the model’s ability to correctly identify true positives and true negatives effectively.

The observed performance differences between the two classifiers can be attributed to SVM’s capacity to handle non-linear relationships in the data. SVM effectively captured intricate patterns within the dataset, leading to enhanced classification accuracy. In contrast, LR’s reliance on linear separation might have limited ability to handle more complex relationships present in the data, resulting in relatively lower performance metrics.

Changes in proteins, lipids, among other compositions of a tissue or biological sample are key markers for discriminating samples from healthy individuals and samples from patients with some type of pathology, such as tissues with the presence of tumor cells. These compositional characteristics can give rise to distinctive characteristics. As reported by Cameron et al.[24], partial gelatinization, crack formation, concentration gradient, and various heterogeneities can be observed in bio-fluid micro-droplets after drying. The drying process, dilution, and environmental conditions impact the pattern formed after drying[24]. Such a pattern becomes important for various applications such as in industrial processes in drug screening, DNA imprinting bioassays, and also in the area of vibrational spectroscopy, especially for the use of body bio-fluids for diagnostics[38].

During microdroplet evaporation, three behaviors associated with specific phase transitions can be identified: sol-gel transition, salt or macro-molecule crystallization, and the crystallization and glass transition in colloidal droplets[39]. Signatures of these processes can be observed in the dried spot, such as the shape and size of crystals formed, the morphology of crystalline aggregates, or the morphology of crack patterns in the residue left after drying[39].

For example, Bahmani, Neysari, and Maleki[40] investigated the formation of patterns of droplets from whole blood samples of normal and thalassemia affected individuals. The authors concluded that the formation of the pattern of fissures is very distinct between the groups and, to explain their results, they invoked the differences in bilirubin level and pressure due to shear viscosity dispersion in blood, volume fraction of particles in close packages and initial film thickness.

Furthermore, substrate choice was found to influence the performance of the classification models. SVM’s higher accuracy on the glass substrate compared to Pt suggests potential variations in sample-substrate interactions. This discrepancy could be attributed to differences in surface properties and their impact on saliva sample behavior on the respective substrates. Our findings emphasize the importance of considering substrate effects and surface interactions when developing classification models for saliva-based diagnostic applications.

## 4 Conclusion

This investigation employed a comprehensive analysis of saliva micro-droplets onto glass and Pt substrates, focusing on OD and SA as key parameters for classification using different mathematical models. LR and SVM models were used to classify the samples into OSCC or healthy groups based on their features. SVM’s use of kernel functions allowed for capturing intricate patterns within the data-set, leading to improved classification accuracy compared to the linear separation approach of LR.

LR models provide valuable insights into how OD and SA predictors influence the likelihood of a sample belonging to a specific group. In contrast, SVM models excel at established optimal decision boundaries that effectively separate the two groups, enabling accurate classification of new, previously unseen samples (see table 1). The two models complement each other, offering a comprehensive understanding of the dataset’s classification patterns. While LR is prized for its interpretability and ease of implementation, SVM handles complex, non-linear relationships between predictors and outcomes through the use of kernel functions.

Significant differences in SA between the glass and Pt substrates could be attributed to varying sample deposition and spreading behaviors on different surfaces, driven by distinct physical properties. The OD differences may also be related to sample composition, thickness, or interactions with the substrates, influencing light scattering or absorption.

These findings pave the way for further advancements in the field, bringing us closer to the development of accurate and reliable diagnostic methodologies using sample imaging setups. The analysis process remains quick, cheap, easily reproducible, and non-invasive for the patients, requiring only 1*µ*l of saliva pipetted onto the substrate for accurate classification as positive or negative for carcinoma. It is important to expand the investigation to other kinds of diseases as infectocontagious (e.g., COVID, influenza and other) and increase the amount of sampled individuals.

## Data Availability

All data produced in the present study are available upon reasonable request to the authors

## Author contributions

G.N.: experimental tests, first draft of the paper, interpretation of data, statistical modeling, revision of the final version of the manuscript. L.F.: sample preparation, microscope data acquisition, experimental tests, statistical modeling, interpretation of data, revision of the final version of the manuscript. M.S.A., N.S.R, and C.M.B.: responsible for sample collection and preparation of the collected saliva. M.G.O.A.: responsible for sample collection and preparation of the collected saliva, and review of the final version of the manuscript. J.T.F.: microscope data acquisition. J.D.A: conceived and designed the experiments for sample collection, and review of the final version of the manuscript. H.S.M.: design of experiments, interpretation of data, and revision of the final version of the manuscript.

## Acknowledgments

The authors are grateful to José Francisco Sales Chaga, Maria Beatriz Nogueira Pascoal and Flávio Francisco Godoy Peres, with the collection of saliva from OSCC patients. They also would like to express their most sincere gratitude to all patients for their time and effort and to thank both the Biotechnoscience and Biophotonics Research Group and the experimental resources provided by Multiuser Central Facilities at UFABC (CEM/UFABC).

This research was funded by the São Paulo Research Foundation (FAPESP) grant #2021/10563-9 and #2016/08633-0; the National Council of Technological and Scientific Development (CNPq): scholarship for MSA; and Coordination for the Improvement of Higher Education Personnel (CAPES): scholarship for MSA and NR.

## Notes

### Competing Interest Statement

The authors have declared no competing interest.

### Funding Statement

This research was funded by the Sao Paulo Research Foundation (FAPESP) grants #2021/10563-9 and #2016/08633-0; National Council for Scientific and Technological Development (CNPq) proc.311933/2021-1, 406761/2022-1 (INCT INTERAS).

### Author Declarations

Research Ethics Committee of Institute of Science and Technology of Sao Paulo State University gave ethical approval for this work.

